# Neural Correlates of Impulsive Compulsive Behaviors in Parkinson’s Disease: A Japanese Retrospective Study

**DOI:** 10.1101/2022.09.15.22280013

**Authors:** Ikko Kimura, Gajanan S. Revankar, Kotaro Ogawa, Kaoru Amano, Yuta Kajiyama, Hideki Mochizuki

**Affiliations:** Department of Neurology, Osaka University Graduate School of Medicine, Suita 565-0871, Japan; Graduate School of Frontier Biosciences, Osaka University, Suita 565-0871, Japan; Graduate School of Information Science and Technology, The University of Tokyo, Tokyo 113-8656, Japan

**Keywords:** Parkinson’s Disease, Impulsive Compulsive Behaviors, Functional Connectivity, Voxel Based Morphometry, Cortico-striatal Network

## Abstract

**Background:** Impulsive compulsive behavior (ICB) often disturb patients with Parkinson’s Disease (PD), of which impulse control disorder (ICD) and dopamine dysregulation syndrome (DDS) are two major subsets. Although the nucleus accumbens (NAcc) is involved in ICB, it remains unclear how the NAcc affects cortical function and defines the different behavioral characteristics of ICD and DDS.

**Objectives:** To identify the most involved cortico-striatal network in ICB and the differences in these networks between patients with ICD and those with DDS using structural and resting-state functional magnetic resonance imaging.

**Methods:** Patients with PD were recruited using the data from a previous cohort study and were divided into patients with ICB (ICB group) and those without ICB (non-ICB group) using the Japanese version of the Questionnaire for Impulsive Compulsive Disorders in Parkinson’s Disease (J-QUIP). From these two groups, we extracted 37 pairs matched for age, sex, disease duration, and levodopa equivalent daily dose of dopamine agonists. Patients with ICB were further classified as either having ICD or DDS based on the J-QUIP subscore. General linear models were used to compare the gray matter volume and functional connectivity (FC) of the NAcc, caudate, and putamen between the ICB and non-ICB groups and between patients with ICD and those with DDS.

**Results:** We found no significant differences in gray matter volume between the ICB and non-ICB groups or between patients with ICD and those with DDS. Compared with the non-ICB group, the FC of the right NAcc in the ICB group was lower in the bilateral ventromedial prefrontal cortex and higher in the left middle occipital gyrus. Patients with DDS showed higher FC between the right putamen and left superior temporal gyrus and higher FC between the left caudate and bilateral middle occipital gyrus than patients with ICD. In contrast, patients with ICD exhibited higher FC between the left NAcc and the right posterior cingulate cortex than patients with DSS.

**Conclusions:** The functionally altered network between the right NAcc and ventromedial prefrontal cortex was associated with the presence of ICB in PD, and the surrounding cortico-striatal networks may differentiate between the behavioral characteristics of patients with ICD and those with DDS.

## 1. Introduction

Parkinson’s disease (PD) is a neurodegenerative disease characterized by the deposition of alpha-synuclein and the degeneration of dopamine-producing cells in the substantia nigra (Rocha et al., 2018). Patients with PD present various motor and non-motor symptoms, including impulsive compulsive behavior (ICB). ICB is the state where patients are unable to resist a certain urge or impulse (Zhang et al., 2014), and 6–34.8% of patients with PD suffer from ICB during the treatment course of dopamine replacement therapy (DRT). While DRT improves motor symptoms, this therapy can exacerbate ICB and no established treatments were there for ICB (Weintraub and Claassen, 2017). Although symptoms associated with ICB worsen the quality of life of the patient and burden of caregivers (Voon et al., 2017; Weintraub and Nirenberg, 2013), ICB are commonly overlooked in clinical practice since patients hesitate to spontaneously report these behaviors owing to shame or denial of their symptoms (Evans et al., 2009; Perez-Lloret et al., 2012). Therefore, objective biomarkers, such as functional connectivity (FC) (Koike et al., 2021) or gray matter volume (GMV) (Kanai and Rees, 2011), are required to detect ICB before these symptoms interfere with patients’ social lives.

The cause of ICB has been attributed to the non-physiological dopaminergic stimulation to the relatively intact regions within the degenerated cortico-striatal networks in PD, including nucleus accumbens (NAcc) (Vriend, 2018).

The deposition of alpha-synuclein in NAcc is lower in patients with ICB than in those without (Barbosa et al., 2019). Moreover, a recent meta-analysis of functional imaging studies revealed that hyperactivation of the NAcc was significantly associated with the presence of ICB (Santangelo et al., 2019). However, there remain no consistent results regarding which alterations in cortico-striatal interactions, including NAcc, lead to ICB. While previous studies revealed alterations in the FC in the fronto-striatal networks of the NAcc in patients with ICB (Mata-Marín et al., 2021; Navalpotro-Gomez et al., 2020; Tessitore et al., 2017), others have suggested the involvement of the FC in other subcortical regions, such as the caudate (Ruitenberg et al., 2018) and putamen (Carriere et al., 2015; Ruitenberg et al., 2018). Previous studies have also shown inconsistent results regarding which regions of the GMV are altered in patients with ICB (Santangelo et al., 2019). One major reason for these inconsistencies is attributed to the relatively small sample sizes in each study (Marek et al., 2022).

Another reason for the inconsistent results on the neural correlates of ICB across studies would be its heterogeneity (Weintraub and Claassen, 2017). ICB comprises two major subsets, namely impulse control disorder (ICD; intolerance of resisting an impulse to perform a certain behavior) and dopamine dysregulation syndrome (DDS; compulsive and excessive use of dopaminergic drugs) (Weintraub et al., 2015). Several case reports have shown that while dopamine agonists (DA) preferentially cause ICD, levodopa causes DDS (Ceravolo et al., 2010; Evans et al., 2009). These reports suggest that different brain regions or mechanisms may be involved in the two symptoms. However, it remains unclear which regions are specifically involved in ICD or DDS development; hence identifying the region specific to each ICB subset is essential for the neuromodulation therapies (Evans et al., 2009).

Therefore, this study identifies the most relevant cortico-striatal networks in the occurrence of ICB and the differences in these networks between patients with ICD and those with DDS. For this purpose, we conducted a retrospective study to compare the FC of cortico-striatal networks or GMV between patients with and without ICB and between patients with ICD and those with DDS. We hypothesize that regions associated with the NAcc are functionally or structurally altered in patients with ICB.

## 2. Material and methods

### 2.1 Participants

This study was conducted as part of a prospective and exploratory study of disease-specific biomarkers and objective indicators of neurodegenerative diseases at Osaka University (UMIN ID: UMIN000036570), which registered all patients with PD admitted to Osaka University Hospital (Kajiyama et al., 2021; Nakano et al., 2021; Otomune et al., 2019). We obtained data on those with or without ICB. We enrolled patients who met the following inclusion criteria: (1) aged 40–85 years, (2) diagnosed with clinically established or probable PD according to the Movement Disorder Society Parkinson Disease Diagnostic Criteria (Postuma et al., 2015), (3) completed resting-state functional magnetic resonance imaging (rsfMRI) and structural MRI scans, and (4) completed the Japanese version of the Questionnaire for Impulsive-Compulsive Disorders in Parkinson’s Disease (J-QUIP). The exclusion criteria were as follows: (1) a history of other neurological or psychiatric diseases and (2) significant neurological abnormalities on MRI scans (e.g., brain tumor or cerebral infarction) evaluated by two neurologists (K.I. and K.Y.). Based on these criteria, 184 patients were included in this study.

### 2.2 Ethics

This study was approved by the Osaka University clinical research review committee (Approval number: 13471-12) and was performed in accordance with the Declaration of Helsinki. Written informed consent was obtained from all participants.

### 2.3 Evaluation for ICB

We divided the eligible patients into two groups namely ICB and non-ICB groups. The ICB group was defined as having a score greater than zero in J-QUIP, whereas the non-ICB group had a score equal to zero (Takeshige-Amano et al., 2022; Tanaka et al., 2013). To investigate the characteristics of ICD and DDS, we further defined patients with ICD as those who responded yes to any item in the following sections of the J-QUIP: A (pathological gambling), B (hypersexuality), C (compulsive buying), and D (binge eating). In contrast, we defined patients with DDS as those who responded yes to any item in section F (DDS) of the J-QUIP.

### 2.4 Clinical evaluations

The following information was obtained from the registered data to acquire the basic characteristics of each group: age, sex, handedness, dominant side of motor symptoms, disease duration, medications, levodopa equivalent daily dose (LEDD), Apathy Scale, Epworth Sleepiness Scale, Frontal Assessment Battery, Geriatric Depression Scale, Hamilton Depression Rating Scale, Mini-Mental State Examination, Parkinson’s Disease Questionnaire-39, and Movement Disorder Society-Sponsored Revision of the Unified Parkinson’s Disease Rating Scale. The LEDD was calculated according to a previous report (Tomlinson et al., 2010).

### 2.5 Image acquisition

MRI data were collected using a GE 3T scanner (GE Medical Systems, WI, USA) in a dark room at Osaka University Hospital. Standard foam pads were placed in the scanner to stabilize the patients’ heads. rsfMRI data were acquired with axial gradient-echo echo-planar imaging (voxel size = 3.3 × 3.3 × 3.2 mm, slice gap = 0.8 mm, matrix size = 64 × 64 × 40, TE = 30 ms, TR = 2.5 s, flip angle = 80°, 240 volumes). During rsfMRI scans, patients were required to keep their eyes fixated on a black cross located at the center of the screen and their bodies as steady as possible, without any thoughts in their minds. Structural MRI data were obtained with a T1-weighted sagittal inversion-recovery spoiled-gradient-echo sequence (voxel size = 1.2 × 1 × 1 mm, matrix size = 200 × 256 × 256, TE = 3.2 ms, TR = 8.2 ms, inversion time = 400 ms). Both MRI scans were collected during the “on” state of each patient.

### 2.6 Quality control for MRI scans

To ensure the image quality of MRI scans, the data were excluded based on the following criteria: (1) significant motion was observed by visual inspection of structural MRI and rsfMRI data (n = 6) and (2) excessive head motion was detected in rsfMRI scans defined as mean framewise displacement (FD) > 0.2 mm, exceeded 20% prevalence of scans with FD > 0.5 mm, or maximum FD > 5 mm (Parkes et al., 2018) (n = 28). We excluded 34 patients, and the data of the remaining 150 patients were used for further analysis.

### 2.7 Image analysis

To conduct FC analysis, T1-weighted and rsfMRI data were preprocessed with the default pipeline of fMRIPrep 20.2.1 (Esteban et al., 2019). T1-weighted data were intensity-normalized, skull-stripped, and tissue-segmented into cerebrospinal fluid (CSF), white matter (WM), and gray matter (GM). These data were then spatially normalized to the Montreal Neurological Institute nonlinear 6th-generation space (MNI standard space). For rsfMRI data, we first skull-stripped the entire dataset, defining the first rsfMRI data as the reference for co-registration to the T1-weighted data. The fMRI data were then corrected for motion and slice timing, warped to the MNI standard space, spatially smoothed with a Gaussian kernel of 6 mm full-width half maximum (FWHM), and temporally band-pass filtered between 0.001 and 0.01 Hz. Several nuisance signals were extracted from the preprocessed rsfMRI data (six head-motion parameters; global signals from the CSF, WM, and the whole brain; motion outliers). We defined the head-motion parameters from the estimates of the motion-correction step and the motion outliers as FD > 0.5 mm or standardized DVARS (Power et al., 2014) > 1.5 standard deviation (Parkes et al., 2018). DVARS was calculated according to a previous study (Power et al., 2014).

To investigate the FC of the NAcc and other striata, namely the caudate and putamen, seed-based correlation analysis was conducted with Nilearn 0.8.1 (https://nilearn.github.io/stable/index.html). Six seeds (the right or left NAcc, putamen, and caudate) were used and defined using Harvard-Oxford cortical and subcortical structural atlases (Desikan et al., 2006). FC between each seed and each voxel in the whole brain was calculated using Fisher’s z-transformed Pearson’s correlation coefficient (z-value) while regressing out the nuisance signals described above.

To analyze GMV, T1-weighted data were preprocessed separately using Statistical Parametric Mapping 12 (SPM12) (Friston, 2003) on MATLAB R2020a (Mathworks, Natick, MA). The details of the preprocessing steps have been described in our previous study (Kajiyama et al., 2021). In brief, T1-weighted data were segmented into CSF, GM, and WM; spatially normalized to the MNI standard space; spatially smoothed with a Gaussian kernel of 8 mm FWHM.

### 2.8 Statistical analysis

To compare the basic and neuroimaging characteristics between the ICB and non-ICB groups, we first extracted pairs from the two groups that were matched for age, sex, disease duration, and LEDD of the DA. Disease duration and DA dose were used for extracting the pairs because both are major risk factors of ICB (Evans et al., 2009) and affect other non-motor symptoms.

For the comparison of basic characteristics between the ICB and non-ICB groups, Mann–Whitney *U* tests were applied for continuous variables. For categorical variables, chi-squared tests were applied, and if the ratio of cells with an expected frequency < 5 was more than 20%, Fisher’s exact tests were applied (Kim, 2017). These tests were performed using R (version 4.1.2, https://www.r-project.org/), and *P* < 0.05 was considered statistically significant.

A second-level general linear model analysis implemented in SPM12 was used to compare FC on each seed or GMV between the ICB and non-ICB groups and between patients with ICD and those with DDS. To compare the ICB and non-ICB groups, we performed two-tailed *t*-tests for FC and an analysis of covariance for GMV, defining total brain volume (TBV) as covariates of no interest. To compare patients with ICD and those with DDS, two separate binary variables (i.e., whether they had ICD/DDS) were first regressed against FC or GMV within the ICB group, setting TBV as covariates of no interest in GMV. We then assessed the differences between the coefficients of ICD (ICD effect) and DDS (DDS effect) using a two-tailed t-test. A voxel-level uncorrected *P* < 0.001 and a cluster-wise family-wise error-corrected *P* < 0.05 were considered statistically significant.

To rule out the possibility of involvement of possible confounders in each of the significant clusters, we calculated Spearman’s correlation coefficients between the mean z-value of each significant cluster and each continuous variable in the basic characteristics. We also performed Mann–Whitney *U* tests on the mean z-value of each significant cluster divided by each categorical variable for the basic characteristics.

### 2.9 Data and code availability

The data for the clinical evaluations and codes used for this analysis are available at github. The raw MRI data are not openly available due to privacy restrictions of clinical data but are available upon a reasonable request from the corresponding authors.

## 3. Results

### 3.1 Basic characteristics

From the 150 eligible patients, we extracted 37 pairs of ICB and non-ICB groups.

According to the J-QUIP, 22, 13, and 8 patients had ICD, DDS, and both, respectively. Between the ICB and non-ICB groups, we found no significant differences in basic characteristics (Table 1; see Supplementary Table 1 and Supplementary Table 2 for the basic characteristics of 150 eligible patients and patients with ICD or DDS, respectively).

**Table 1.**
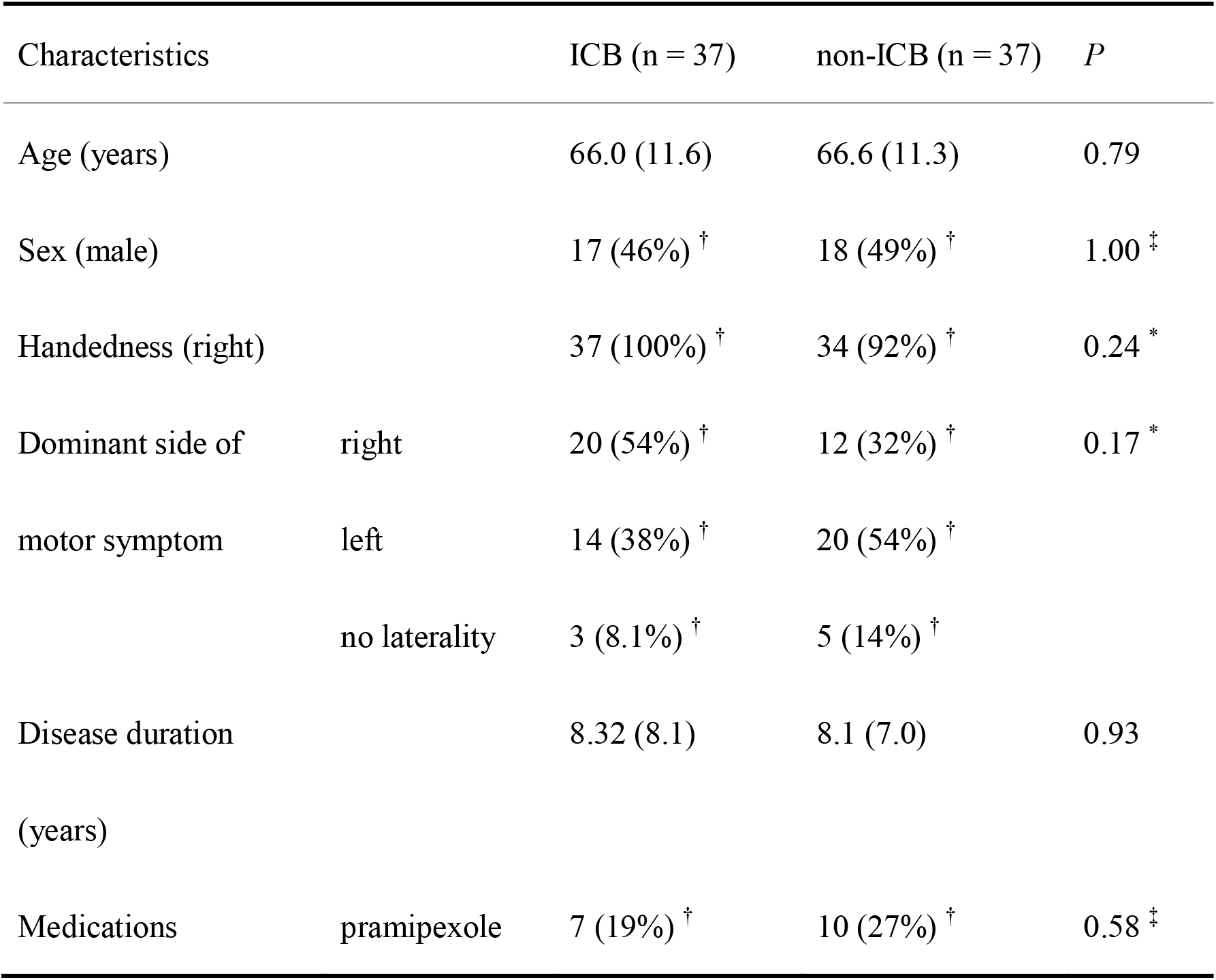
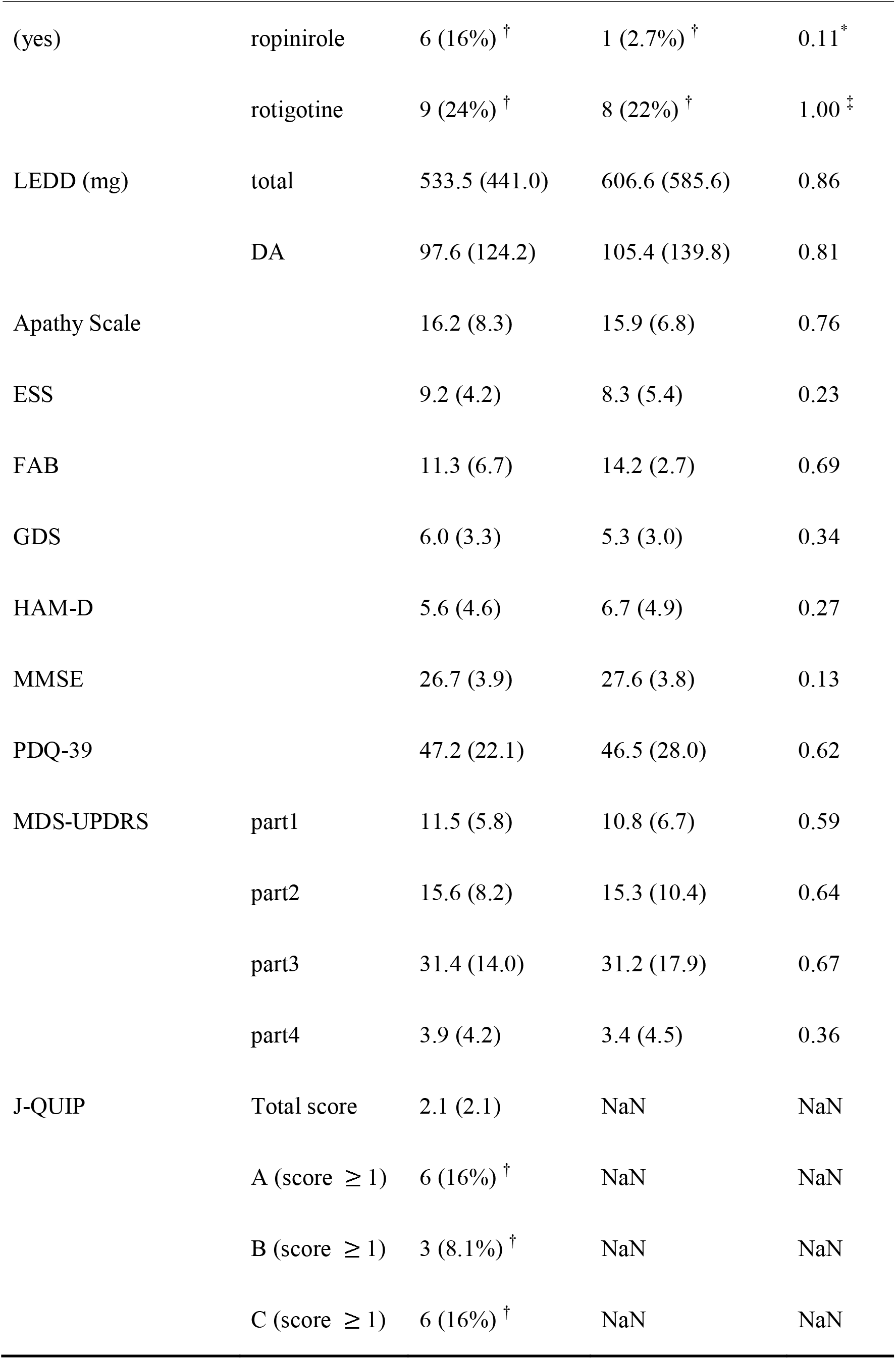
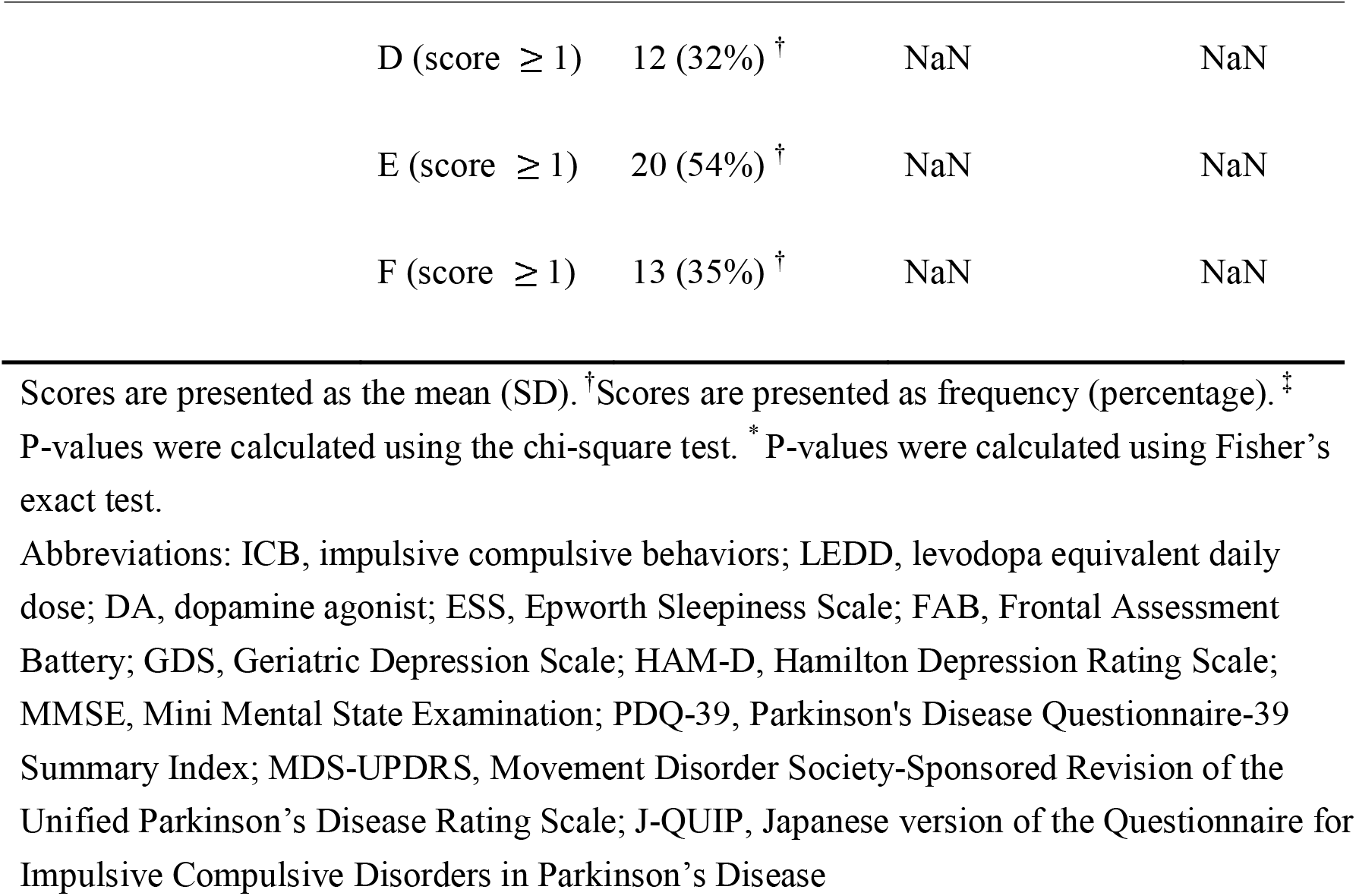
Basic characteristics of patients with Parkinson’s disease in the ICB and non-ICB group

### 3.2 Comparisons between the ICB and non-ICB groups

We first compared FC for six seeds (the right or left NAcc, putamen and caudate) between ICB and non-ICB groups. Comparisons concerning the FC of the right NAcc revealed that the FC of the ICB group was significantly lower in the bilateral ventromedial prefrontal cortex (vmPFC) than that of the non-ICB group (Figure 1A; see Supplementary Table 3 for statistical values and peak MNI-coordinates on each significant cluster). The mean z-values of this significant cluster were 0.10 ± 0.10 and 0.25 ± 0.12 in the ICB and non-ICB groups, respectively (Figure 1B). In contrast, the FC of the ICB group was significantly higher in the left middle occipital gyrus (MOG) than that of the non-ICB group (Figure 1A). The mean z-values of FC in this cluster were 0.019 ± 0.083 and -0.090 ± 0.088 in the ICB and non-ICB groups, respectively (Figure 1C). We observed no significant differences between the ICB and non-ICB groups regarding the FC in the left NAcc, bilateral caudate, or bilateral putamen and the GMV.

**Figure 1.**
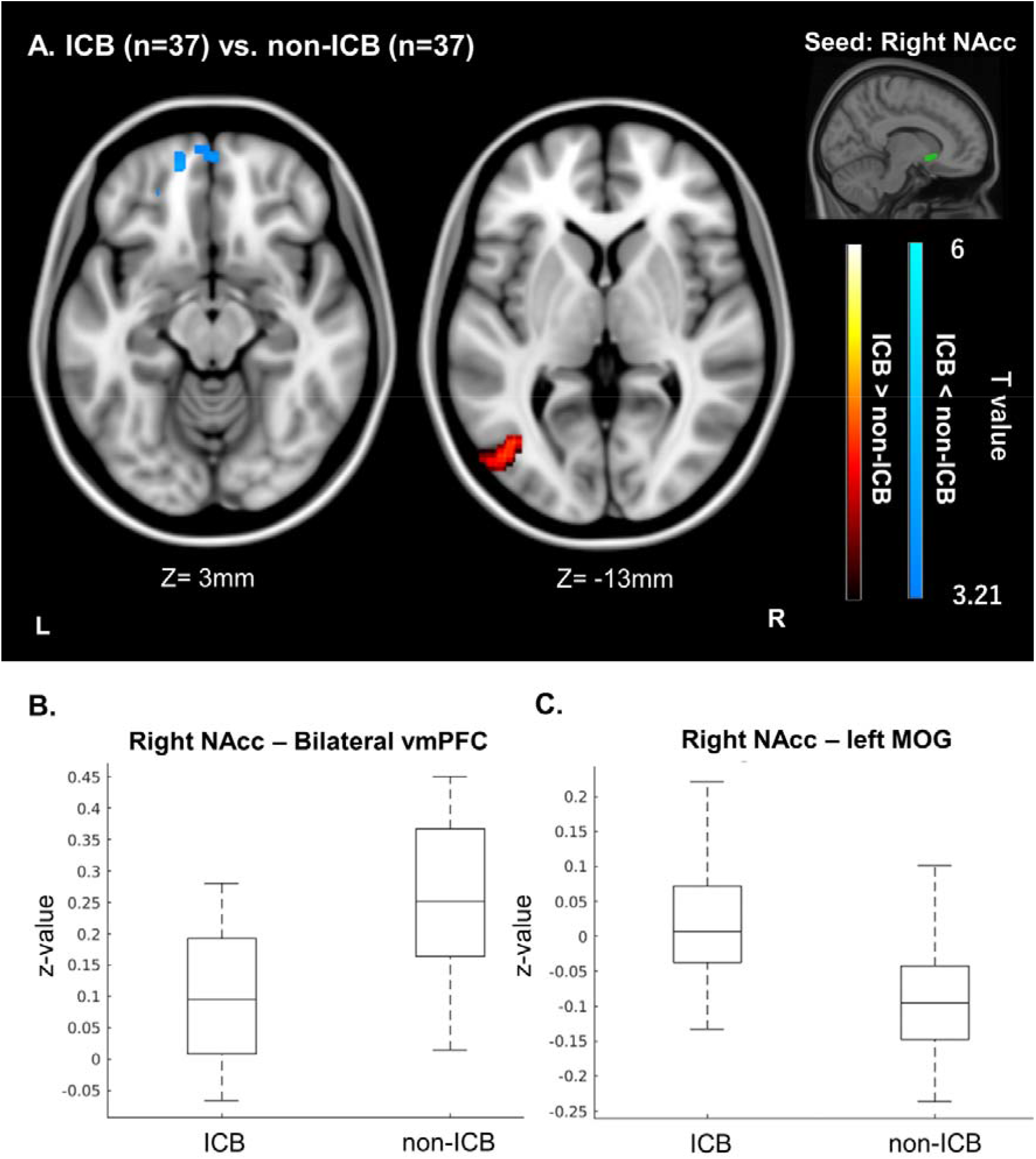
Seed-based correlation analysis comparing the functional connectivity between the ICB and non-ICB groups. (A) The resulting spatial maps. The green shaded region indicates the seed (right NAcc). The axial slices are displayed according to the neurological convention (left on the picture is left in the brain). (B, C) Boxplot of the mean Fisher-transformed Pearson’s correlation coefficients (z-values) of each significant cluster in each group. Abbreviations: ICB, impulsive compulsive behaviors; NAcc, nucleus accumbens; vmPFC, ventromedial prefrontal cortex; MOG, middle occipital gyrus

### 3.3 Comparisons between the ICD and DDS effects

We then compared FC and GMV between patients with ICD and those with DDS and found that the FC of patients with ICD was lower between the right putamen and left superior temporal gyrus (STG) and between the left caudate and bilateral MOG than that of patients with DDS (Figure 2A). The mean z-values on the FC between the right putamen and left STG were -0.041 ± 0.18, 0.12 ± 0.13, and 0.019 ± 0.12 in patients with ICD, those with DDS, and the non-ICB group, respectively (Figure 2B). The mean z-values on the FC between the left caudate and bilateral MOG were -0.13 ± 0.089, 0.0025 ± 0.11, and -0.11 ± 0.093 in patients with ICD, those with DDS, and the non-ICB group, respectively (Figure 2C). In contrast, the FC between the left NAcc and right posterior cingulate cortex (PCC) in patients with ICD was lower than that in patients with DDS (Figure 2A). The mean z-values of the FC between the left NAcc and right PCC were 0.055 ± 0.12, -0.082 ± 0.10, and 0.029 ± 0.098 in patients with ICD, those with DDS, and the non-ICB group, respectively (Figure 2D). We found no significant differences between patients with ICD and those with DDS regarding the FC of the right caudate, right NAcc, or left putamen and the GMV.

**Figure 2.**
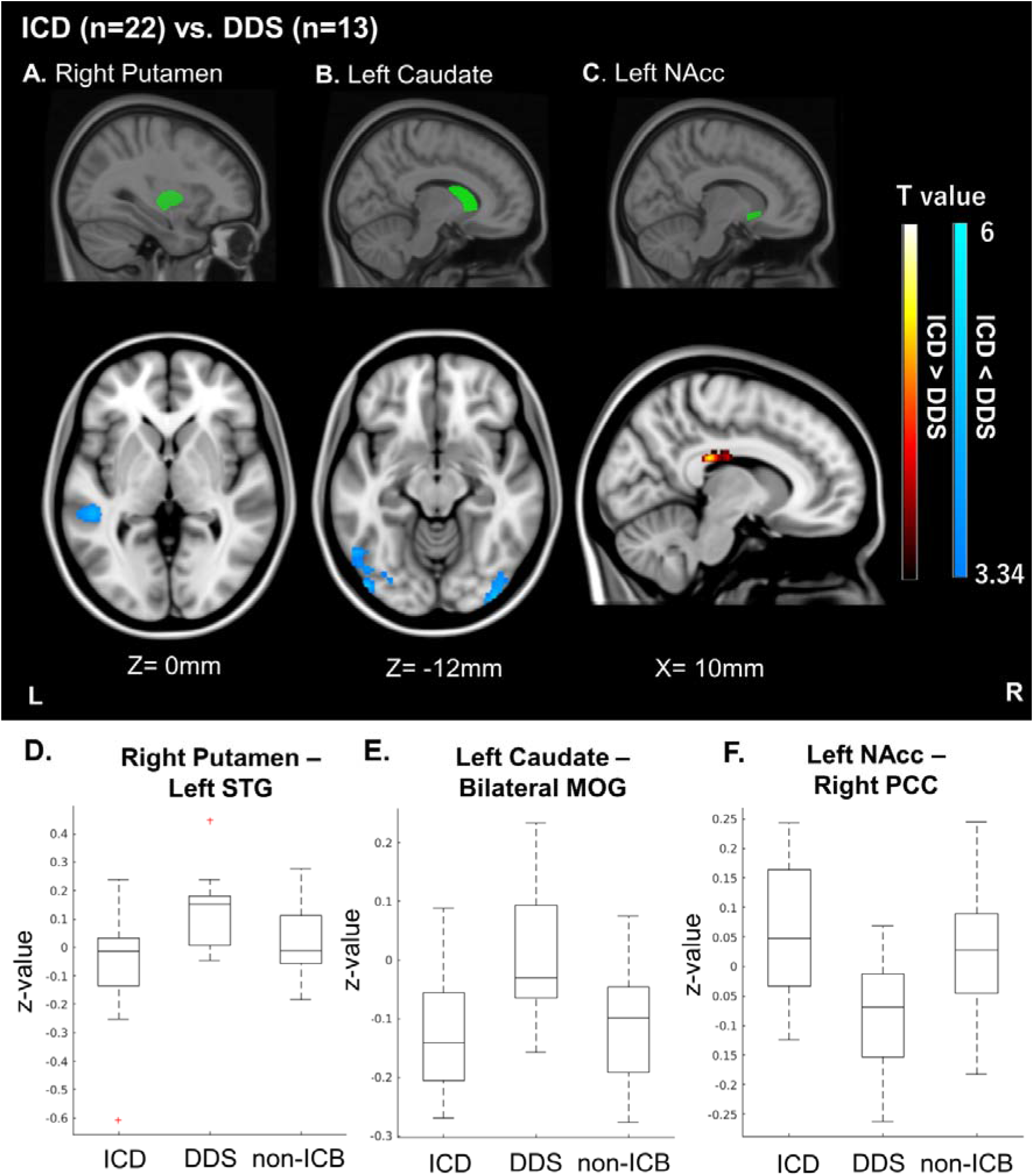
Seed-based correlation analysis comparing the functional connectivity between the ICD and DDS effects in patients with PD. The resulting spatial maps using (A) right putamen, (B) left caudate, and (C) left NAcc as a seed. The green shaded regions indicate the seed. The axial slices are displayed according to the neurological convention (left on the picture is left in the brain). (D – F) Boxplots of the mean z-value of each significant cluster in patients with ICD, with DDS, and without ICB, respectively. A red-cross indicates an outlier detected by the default settings of “boxplot” in MATLAB. Abbreviations: ICD, impulse control disorder; DDS, dopamine dysregulation syndrome; ICB, impulsive compulsive behaviors; PD, Parkinson’s disease; STG, superior temporal gyrus; MOG, middle occipital gyrus; NAcc, nucleus accumbens; PCC, posterior cingulate cortex

### 3.4 Dependencies with possible confounding factors

We found no significant dependencies of the basic characteristics on the mean z-value of each significant cluster when comparing between the ICB and non-ICB groups (Supplementary Table 4) or between ICD and DDS effects (Supplementary Table 5).

## 4. Discussion

To the best of our knowledge, our study included the largest sample size compared with other related previous studies (Santangelo et al., 2019). Here, we found that the FC of the right NAcc differed between the ICB and non-ICB groups and that the FC of the left NAcc, left caudate, and right putamen were altered in patients with ICD and those with DDS. None of these significant clusters was dependent on any other clinical demographics, suggesting that these differences were specifically derived from differences in ICB characteristics.

### 4.1 FC of the right NAcc was altered in the ICB group

The FC between the right NAcc and bilateral vmPFC was lower in the ICB group than in the non-ICB group, whereas the FC between the right NAcc and left MOG was higher in the ICB group. Several neuroimaging studies have revealed that the activity of the NAcc was increased in patients with ICB (Santangelo et al., 2019). One study reported that fluorodopa uptake in patients with ICB increased in the medial regions of the prefrontal cortex (Joutsa et al., 2012). These results, including ours, suggest that both the NAcc and vmPFC are functionally altered in patients with ICB, which is consistent with our finding that the FC between these two regions decreased. Moreover, the vmPFC is structurally and functionally linked to the NAcc through the direct projections of glutamate neurons (Rusche et al., 2021; Wichmann and Delong, 2006). Previous neuroimaging studies have shown that while the NAcc has a broad function in reward-based learning, the vmPFC plays a specific role in evaluating reward value (Haber and Knutson, 2010; Liu et al., 2011). Furthermore, several studies have reported that the vmPFC is crucial for risky decision-making (Clark, 2010; Rogalsky et al., 2012). Therefore, a decrease in the FC between these regions might disrupt reward-based learning, which in turn induces ICB.

In contrast, another study reported the disruption of visuospatial memory in patients with ICB (Vitale et al., 2011). Our results also reflect this disruption by showing an increase in FC between the right NAcc and left MOG since the MOG plays a critical role in visual processing (Wang et al., 2015). We acknowledge that the NAcc and MOG are not directly connected, but indirectly connected via thalamus (Takeshige-Amano et al., 2022), and this increase of FC might reflect the disruption of other cognitive functions such as visual emotion recognition (Takeshige-Amano et al., 2022). Thus, we can only speculate on the functional role of the increase in FC between these two regions in the occurrence of ICB in patients with PD. Further neuropsychological studies are required to clarify this role.

No significant difference in the FC between the putamen and caudate was found. The reason for this specificity might be that the NAcc in patients with ICB was relatively intact compared with other subregions of the striatum. The load of alpha-synuclein in the NAcc was lower in patients with ICB than in those without ICB, whereas that in the putamen or caudate was not significantly different (Barbosa et al., 2019). Furthermore, the ventral striatum was relatively intact compared with other regions of the striatum until the late stage of PD (Cools, 2006). Taken together, the NAcc may contribute more to the occurrence of ICB than the putamen or caudate.

We found a significant difference in the FC of the right NAcc but not in that of the left NAcc. The right NAcc might be more important in controlling reward-based learning than the left NAcc as several studies have shown that action inhibition is right-dominant (Aron et al., 2004; Garavan et al., 1999; Rubia et al., 2003). Another reason for this laterality may be the rather high rate of right dominance of motor symptoms in the ICB group, although the rate of the dominant side of motor symptoms was not significantly different between the ICB and non-ICB groups. In fact, the right-sided onset of motor symptoms has recently been proposed to be another risk factor for ICB (Phillipps et al., 2020). The right dominance of motor symptoms means that the right striatum is intact compared with the left striatum, which might cause an overdose of dopamine in the right striatum. Therefore, the right NAcc is the major component that induces ICB in patients with PD.

In contrast, two studies reported significant alterations in the FC of the caudate and putamen in patients with ICB (Carriere et al., 2015; Ruitenberg et al., 2018), while other studies have suggested the involvement of the dorsal striatum in reward-based learning (Haber and Knutson, 2010). These differences might be due to the different definitions of seeds or distributions of subcategories in the ICB (e.g., The rate of hypersexuality in studies by Carrire et al. (2015) and Ruitenberg et al. (2018) was 70% and 43%, respectively, while the rate in our study was 8.1%). Therefore, different cortico-striatal networks may be associated with different ICB subcategories. Future studies are required to clarify the neural correlates of each ICB subcategory.

### 4.2 FC of striatum was different in patients with ICD and those with DDS

We observed a decrease in FC between the left NAcc and right PCC in patients with DDS compared with those with ICD. The PCC and NAcc play critical roles in reward-based learning (Liu et al., 2011). The decrease in FC in these regions reflects that the reward-processing steps are differentially altered in patients with ICD and DDS. Another reason for the decrease in FC between the right PCC and left NAcc in patients with DDS may be the difference in the sensitivity to levodopa between patients with ICD and those with DDS. Several case reports have indicated that levodopa preferentially induces DDS (Evans et al., 2009). One study also reported that levodopa decreased the FC between the ventral striatum and PCC (Kelly et al., 2009). Therefore, patients with DDS might be more sensitive to levodopa than those with ICD.

We also found an increase in FC between the right putamen and left STG and between the left caudate and bilateral MOG in patients with DDS. The left STG is crucial for semantic processing (Price, 2012), whereas MOG is important for visual processing (Wang et al., 2015). Taken together, the involvement of the striatum in semantic or visual processing may differ between patients with ICD and DDS. Therefore, the functional relationship between the striatum and regions related to reward, semantic, or visual processing might be differentially altered in patients with ICD compared with those with DDS. These altered neural circuits may explain ICB heterogeneity.

### 4.3 No significant difference in the GMV

While we found significant differences in the FC of the striatum according to ICB features, we detected no significant differences in the GMV. Four out of ten previous studies comparing patients with and without ICB showed non-significant differences in GMV between the ICB and non-ICB groups (Santangelo et al., 2019). ICB are reversible symptoms, since these behaviors can be ameliorated by reducing the dose of DA (Evans et al., 2009). This implies that reversible functional alterations can induce ICB, and our results suggest that functional differences can occur in patients with ICB, even without structural differences. Taken together, functional measurements are more useful than structural measurements for detecting ICB, ICD, and DDS.

### 4.4 Limitations

This study has several limitations. First, we defined ICB using only the J-QUIP. The accuracy of J-QUIP in detecting ICB was comparable to that of an internationally established questionnaire for detecting ICB (Tanaka et al., 2013). Nevertheless, these scores are subjective and may underestimate the status of ICB (Perez-Lloret et al., 2012). Future studies are required to evaluate the status of ICB using objective neuropsychological tests such as the Iowa Gambling Task (Buelow and Suhr, 2009).

Second, this was a retrospective study; therefore, it is not clear whether alterations in FC were induced by DRT or whether they already existed before DRT. Future prospective and longitudinal studies are required to reveal the causality of changes in FC in these significantly different clusters.

Third, the number of patients within some ICB subcategories, such as pathological gambling or hypersexuality, was relatively small. Therefore, we could not differentiate the neural correlates of each ICB subcategory. Future multi-center studies are required to increase the sample size of patients with ICB and to detect the unique neural characteristics of each ICB subcategory.

### 4.5 Conclusions

Our findings showed that cortico-striatal networks play an important role in the occurrence of ICB and that these networks reflect the different characteristics of ICD and DDS. Evaluating the FC of these networks might be useful for detecting ICB, which may be underestimated through clinical interviews alone. Elucidating the altered regions is important not only for identifying biomarkers for ICB but also for clarifying the treatment target of ICB with neuromodulation.

## Data Availability

All data produced in the present study are available upon reasonable request to the authors

## ^1^ Abbreviations

PD: Parkinson’s disease
ICB: impulsive compulsive behaviors
ICD: impulse control disorder
DDS: dopamine dysregulation syndrome
FC: functional connectivity
GMV: gray matter volume
NAcc: nucleus accumbens
J-QUIP: Japanese version of the Questionnaire for Impulsive Compulsive Disorders in Parkinson’s Disease
DRT: dopamine replacement therapy
DA: dopamine agonists
rsfMRI: resting-state functional magnetic resonance imaging
LEDD: levodopa equivalent daily dose
CSF: cerebrospinal fluid
WM: white matter
GM: gray matter
FWHM: full-width half maximum
FD: framewise displacement
TBV: total brain volume
vmPFC: ventromedial prefrontal cortex
MOG: middle occipital gyrus
STG: superior temporal gyrus
PCC: posterior cingulate cortex

## Acknowledgements

This study was supported by Grants-in-Aid from the Research Committee of Central Nervous System Degenerative Diseases, Research on Policy Planning and Evaluation for Rare and Intractable Diseases, Health, Labor and Welfare Sciences Research Grants, the Ministry of Health, Labor and Welfare, Japan (Grant number: 20FC104).

## CRediT author statement

IK: Conceptualization, Methodology, Software, Data Curation, Formal analysis, Writing - Original Draft, Visualization

GR: Writing - Review & Editing

KO: Investigation, Resources, Project Administration

KA: Writing - Review & Editing

YK: Investigation, Resources, Project Administration, Writing - Review & Editing

HM: Supervision, Writing - Review & Editing, Funding Acquisition

